# Algorithmic Multi-Domain Syndromic Profiling in Cerebrovascular Disease: A Deterministic Approach to Post-Stroke Neurocognitive Deficit Mapping

**DOI:** 10.64898/2026.09.08.26362502

**Authors:** Alexander I. Erzin

**Author notes:** **Corresponding Author:** Alexander I. Erzin, Cognicore Neurosystems, WY, USA.

## Abstract

**Background:** Post-assessment data processing, quantitative syndromic profiling, and clinical report generation in acute stroke settings typically require up to 50 minutes of manual documentation per patient, straining clinical workflows and driving clinician burnout. Conversely, standard automated screening tools (NIHSS, MoCA) lack sensitivity to the qualitative specificity of focal deficits and fail to generate structured, auditable clinical narratives.

**Methods:** We performed a retrospective observational study of a prospective acute registry (n = 169; 84 males, 85 females; mean age 66.87 years, SD = 13.9, range 18–97). Cognitive mapping was executed via the NeuroDraft platform, which pairs a deterministic Python core implementing a 10-domain “Luria Raw” matrix (scaled 0–5) with a constrained generative linguistic layer for structured reporting.

**Results:** The deterministic scoring pipeline demonstrated high internal consistency (Cronbach’s α = 0.85). Kruskal-Wallis testing (df = 7, ties-corrected) revealed marked discriminant validity across baseline mental status tiers (p < 0.001), led by visual object perception (H = 44.86) and complex attention (H = 42.55). Principal component analysis of the covariance matrix identified a primary general deficit factor (λ_1_ = 4.13, explaining 41.26% of total variance) dominated by visuoconstructive functions (loading = 0.46), with basic numerical calculation showing relative independence (loading = 0.21). Depressive symptom loading demonstrated negative orthogonality to organic impairment severity (r = -0.20).

**Conclusion:** The NeuroDraft software standardizes and quantifies qualitative Luria’s syndromic analysis, mitigating human-error bias and reducing the medical reporting cycle to 5–10 minutes. This deterministic method provides reproducible cognitive profiling suitable for routine clinical workflows.

## INTRODUCTION

### Clinical Crisis in Acute Stroke Neuropsychological Assessment

Acute ischemic stroke—particularly following middle cerebral artery (MCA) occlusion—triggers a cascading, multidomain breakdown of higher cortical functions. This disruption presents clinically as complex comorbid syndromes of aphasia, apraxia, agnosia, and executive dysfunction (Tsvetkova, 1988). Prompt topical localization and syndromic profiling of this impairment are critical for predicting rehabilitation potential; however, emergency neurology faces fundamental methodological constraints. Widely used screening batteries – such as the National Institutes of Health Stroke Scale (NIHSS), Montreal Cognitive Assessment (MoCA), and Mini-Mental State Examination (MMSE) – exhibit pronounced ceiling and floor effects. As purely quantitative tools, they remain blind to qualitative error patterns (Boston Process Approach). Consequently, they fail to dissociate primary focal cortical defects from generalized fluctuations in central nervous system arousal, including stupor, delirium, or pathological somnolence (Kaplan, 1988; Mikadze et al., 2018). Overcoming these limitations historically relies on gold standards: A.R. Luria’s syndromic analysis and the Boston Process Approach, both designed to isolate the primary underlying deficit (Ashendorf et al., 2013; Glozman, 1999; Poreh, 2000). However, deploying these methodologies into acute hospital workflows introduces a major operational bottleneck: manual administration, scoring, qualitative interpretation, and report writing require 50 to 90 minutes. In high-volume emergency inpatient settings, this manual burden drives clinician burnout, severely constraining clinical throughput and compromising serial dynamic monitoring. This creates an urgent clinical need for tools that digitize qualitative syndromic profiling without sacrificing semiotic depth, mapping quantitative profiles directly onto standardized International Classification of Functioning, Disability and Health (ICF) metrics (World Health Organization, 2001) to facilitate standardized functional monitoring.

### Paradigm Shift: From Narrow Localization to Large-Scale Neural Networks

Modern clinical neuropsychology is undergoing a paradigm shift, bridging Luria’s three functional brain units (Luria, 1973) and systemic dynamic localization models (Anokhin, 1974; Vygotsky, 1965) with contemporary connectomics and large-scale brain networks. Within this architecture, Luria’s subcortical arousal loop (Unit I), posterior sensory processing zone (Unit II), and anterior executive system (Unit III) function as macroeconomic baselines that modulate and sustain large-scale intrinsic connectivity networks, specifically the Default Mode (DMN), Executive Control (ECN), and Salience (SN) networks, whose topologies are well-delineated via resting-state fMRI and graph-theoretical analysis (Bressler & Menon, 2010; Bullmore & Sporns, 2009; Fox et al., 2005; Yeo et al., 2011).

This integrated framework, operationalized across validated international batteries (Peña-Casanova et al., 2006), establishes a direct mechanistic continuum between vascular pathophysiology and neuropsychological profiles. Under this framework, clinicians can dissociate acute focal cortical disruptions (hallmarks of MCA infarctions) from latent subcortical degradation of distributed topological hubs (van den Heuvel & Sporns, 2013) characteristic of cerebral small vessel disease (CSVD). Small vessel disease markers and TOAST stroke subtypes disrupt thalamocortical, reticular, and striatal loops, yielding syndromic patterns distinct from pure cortical infarctions. Furthermore, diagnostic models must accommodate cerebellar cognitive affective syndrome (CCAS / cerebellar dysmetria), which impairs working memory, verbal fluency, and visuospatial cognition despite an intact cerebral cortex (Schmahmann & Sherman, 1998). Recent fMRI studies and clinical guidelines confirm discrete cognitive topographies within the posterior cerebellar lobe functioning as predictive nodes for adaptive control (Guell et al., 2018; Hoche et al., 2018; Sokolov et al., 2017). The characterization of these complex phenotypes warrants models that synthesize cognitive profiles with neuroimaging patterns and angiographic modifiers.

### Empirical Basis and Deterministic Architecture of the NeuroDraft Platform

To address this clinical bottleneck, this study presents the architectural framework and empirical validation of the NeuroDraft software platform. The complete digital pipeline—from bedside assessment data ingestion to the generation of a structured clinical report—operates within 5 to 10 minutes client-side. The empirical foundation for the computational core comprises a prospective hospital registry containing n = 169 verified clinical records collected in a tertiary emergency center. The cohort spans eight acute departments (two acute stroke neurology units, two neurosurgical units, two cardiology units, internal medicine, and traumatology), ensuring representative clinical heterogeneity. Demographic analysis (mean age 66.87 years, range 18–97 years) demonstrated equal biological sex distribution (n = 84 men, n = 85 women), fully adhering to the SAGER guidelines.

The system uses a two-tier deterministic pipeline designed to eliminate stochastic errors:

1. Tier 1 (Deterministic Measurement Core Engine): Implemented in Python, this layer processes 10 continuous fractional domains from the “Luria Raw” matrix (scaled 0 to 5 points). In tandem with angiographic modifiers (*mri_status, clinical_pool*, TOAST subtypes), the engine automatically maps deficit severity onto standardized International Classification of Functioning, Disability and Health (ICF) categories across codes **b** (body functions) and **d** (activities and participation), while algorithmically deriving predictive environmental modifications under codes **e** (environmental factors).
2. Tier 2 (Linguistic Synthesis Layer): A hardware-agnostic, third-party large language model (LLM) integrated via an API gateway, operating strictly as a text-formatting utility within the closed context of the preprocessed deterministic outputs. Because the linguistic layer serves strictly to synthesize structured psychometric outputs into cohesive clinical narratives, the underlying measurement parameters remain independent of the specific large language model choice and do not affect the mathematical validity of the underlying Luria Raw methodology.

Psychometric validation of the deterministic core engine confirmed high internal consistency across the cohort (Cronbach’s α = 0.85). Nonparametric Kruskal-Wallis H-testing (SciPy library), corrected for tied ranks, demonstrated robust discriminant validity across all 10 evaluation domains (p < 0.001). Latent structure extraction via singular value decomposition (PCA / Power Iteration) revealed that the leading eigenvector of the covariance matrix (PC1) was dominated by visuoconstructive functions (D6), which yielded the highest component loading (0.46). Conversely, numerical calculation (D7) contributed the lowest relative loading to this general deficit factor (0.21).

In summary, this approach transforms qualitative bedside assessment into a quantitative computational framework. It delivers a robust, mathematically grounded, and hardware-agnostic instrument for profiling post-stroke aphasias and apraxias during the hyperacute and acute phases of cerebrovascular accidents.

## MATERIALS AND METHODS

### Registry Profile and Data Collection Context (Bedside Context)

This study is a retrospective observational analysis of a prospectively compiled hospital registry comprising n = 169 verified clinical cases. The cohort spans patients with a mean age of 66.87 years (range: 18–97 years). Strict biological sex balance was maintained (n = 84 men, n = 85 women), adhering to the Sex and Gender Equity in Research (SAGER) guidelines.

**Table A.** Nosological and Demographic Characteristics of the Cohort (*n* = 169)

| Clinical Parameter / Modifier | Absolute Count (n) | Percentage (%) |
| --- | --- | --- |
| **Total Cohort Size** | **169** | 100.0% |
| **Biological Sex (SAGER Guidelines):** |  |  |
| • Men (male) | 84 | 49.7% |
| • Women (female) | 85 | 50.3% |
| **Neuroimaging Status (*mri_status*):** |  |  |
| • Code 0 (Normal / No focal lesions) | 45 | 26.6% |
| • Code 1 (Verified structural lesion) | 68 | 40.2% |
| • Code 2 (Acute CT-negative phase) | 56 | 33.1% |
| **Clinical Vascular Territory (*clinical_pool*):** |  |  |
| • Code 0 (Non-vascular / Somatic control) | 55 | 32.5% |
| • Code 1 (Anterior / Carotid circulation) | 74 | 43.8% |
| • Code 2 (Posterior / Vertebrobasilar circulation) | 40 | 23.7% |
| **TOAST Pathogenetic Subtype (*toast_subtype*):** |  |  |
| • Code 1 (LAA – Large-artery atherosclerosis) | 12 | 7.1% |
| • Code 2 (CE – Cardioembolism) | 11 | 6.5% |
| • Code 3 (SVO – Small-vessel occlusion / Lacunar) | 14 | 8.3% |
| • Code 4 (ODE – Other determined etiology) | 16 | 9.5% |
| • Code 5 (UD – Undetermined etiology) | 61 | 36.1% |
| • Code 0 (Not applicable / Somatic control) | 55 | 32.5% |

An expert clinical neuropsychologist performed all primary neuropsychological and clinical assessments directly at the bedside, avoiding the artificial constraints of outpatient testing suites. Assessments took place within acute dedicated stroke units. The testing protocol was tailored for acute bedside settings, accommodating severe hemiparesis, post-stroke fatigue, somatic instability, and motor deficits. This bedside assessment design captured real-time fluctuations in cognitive and behavioral symptoms during the hyperacute and acute phases of cerebrovascular accidents. The registry’s nosological structure captures broad clinical heterogeneity codified via angiographic modifiers. The cohort includes ischemic events across both carotid (pool = 1) and vertebrobasilar (pool = 2) territories. Structural neuroimaging (CT/MRI) verified lesion status, classifying cases into confirmed lesions (mri_status = 1) and acute CT-negative presentations (mri_status = 2). The cohort also includes small vessel disease (SVD) patterns and lacunar strokes classified under TOAST criteria, enabling differential assessment of focal cortical versus diffuse subcortical deficits. Selection Criteria & Inclusion/Exclusion Protocol

Patient selection followed clinical necessity principles and Medicare/CMS Medical Necessity Guidelines for specialist diagnostic procedures.

#### 1. Strict Exclusion Criteria

To isolate pure cerebrovascular pathophysiology, screening excluded:

- Patients with verified primary or metastatic central nervous system (CNS) malignancies.
- Patients with severe isolated or polytraumatic traumatic brain injury (TBI) in the acute or subacute phase.
- Patients with active neuroinflammatory or infectious CNS diseases (encephalitis, meningitis, autoimmune vasculitis).
- Patients with multiple prior ipsilateral or bilateral strokes resulting in extensive cystic-gliotic transformations and severe premorbid disability.

#### 2. Inclusion Criteria and Baseline Control Structure

The primary cohort included patients presenting with first-ever focal ischemic events in the carotid or vertebrobasilar territories. A somatic baseline control group comprised non-stroke cardiology, internal medicine, and orthopedic trauma inpatients. Control subjects had no acute focal CNS lesions, maintained intact verbal communication, and completed all test paradigms without organic cognitive impairment. In accordance with AHA/ASA clinical guidelines, expanded neurocognitive screening in this medical cohort (specifically in acute myocardial infarction and decompensated heart failure) objectively profiles systemic vascular burden. The NeuroDraft platform captures baseline mental status across *k* = 8 discrete levels of the mental_status_code variable, indexing verbal rapport, cooperation, and affective/arousal state upon entry: standard post-stroke profile (Code 0), moderate rapport deficit (Code 1), severe executive-behavioral dysfunction (Codes 2, 3, and 4), complete communication block (Code 5), somatic control emotional lability (Code 7), and transient neurodynamic slowing due to sedation or sleep deprivation (Code 0sleep). This stratification detects cardiogenic encephalopathy and subclinical cognitive decline induced by systemic circulatory hypoxia and cerebral hypoperfusion. This stratification evaluates the algorithms’ discriminant validity in separating acute cerebrovascular events from comorbid baseline decline, which maximizes ecological validity.

#### 3. Subclinical Neurodegenerative Overlay

To validate the sensitivity and flexibility of the NeuroDraft deterministic measurement pipeline, the registry included patients with suspected, indeterminate, or early-stage neurodegenerative diseases (primarily early Parkinson’s disease and early mixed/microangiopathic dementia), representing 1.8% of the cohort. Including this subclinical degenerative cohort tested whether the 10 domains of the Luria Raw matrix can isolate primary focal stroke syndromes (agnosias, aphasias, apraxias) from background cognitive-affective overlay and frontal-subcortical decline typical of synucleinopathies. In suspected Parkinson’s disease cases, NeuroDraft isolated specific decrements in processing speed (complex_attention_speed) and executive function (executive_control_reasoning) with preserved primary cortical gnostic zones, consistent with Movement Disorder Society (MDS) criteria. Testing this controlled 1.8% neurodegenerative overlay demonstrated that the two-tier deterministic architecture successfully discriminates acute focal infarction (carotid/vertebrobasilar stroke) from insidious synaptic breakdown, ensuring diagnostic robustness in polymorbid acute settings. Diagnostic Instrumentation and Subtest Specifications (Luria Raw Engine)

In contrast to reductionist screening tools, this study implemented a comprehensive clinical battery adapting classical tasks from A.R. Luria and N.K. Korsakova (modified by E.Y. Balashova et al.) for rapid acute bedside mapping. The evaluation architecture comprises 10 continuous fractional domains within the “Luria Raw” matrix, scored continuously from 0 (unimpaired/normal) to 5 (complete functional collapse):

- Complex Attention & Processing Speed: Evaluates visual attentional selectivity, vigilance, and set-shifting, alongside processing speed, using adapted visual search matrices and serial subtraction paradigms.
- Visual Object Perception: Measures visual object recognition across canonical representations and degraded stimuli (Poppelreuter overlapping figures, crossed-out drawings, and fragmented contours).
- Visuospatial Processing & Orientation: Tests visuospatial gnosis and mental transformation operations using mental rotation tasks, unnumbered (“blind”) clock face reading, and rapid assessment of topographical orientation.
- Praxis & Motor Sequencing: Assesses motor fluency, kinetic shifting, and automated efferent motor sequences via reciprocal bimanual coordination and the Luria dynamic kinetic sequencing test (fist-edge-palm).
- Kinesthetic & Somatosensory Praxis: Evaluates somatosensory afferent praxis through blind replication of articulatory postures and manual finger configurations, alongside somatosensory spatial localization accuracy.
- Visuoconstructive Functions: Measures visuoconstructive praxis and spatial synthesis via bedside-adapted block design synthesis and geometric pattern replication.
- Numerical Processing & Calculation: Assesses retention of numerical positional syntax, spatial digit alignment, and mental numerical calculation across automated and non-automated sequences.
- Language / Speech: Profiles receptive and expressive language, including minimal-pair phonemic discrimination, confrontation naming, semantic density, and comprehension of inverted logico-grammatical constructions.
- Learning and Memory: Assesses audioverbal and visual mnestic functions, quantifying immediate recall, delayed retention, and learning curves under homogeneous and heterogeneous interference paradigms (short-term and long-term memory).
- Executive Functioning & Conceptual Reasoning: Evaluates supervisory cognitive control, abstraction, and set-shifting using conceptual categorization, similarities, odd-one-out paradigms, and figurative proverb/metaphor interpretation.

### Information Security Framework (Zero-Knowledge Architecture)

To uphold strict patient confidentiality, the platform employs a zero-knowledge cryptographic architecture. All personally identifiable information (PII) processing and raw text transcription execute client-side. The computational server receives only anonymized raw numeric tensors. This routing framework ensures full legal compliance with HIPAA and GDPR standards.

### Mathematical and Statistical Processing Pipeline (Statistical Pipeline)

All primary psychometric calculations are executed via a deterministic Python architecture, using standard linear algebra libraries and SciPy (scipy.stats) to preclude stochastic errors during the diagnostic scoring phase. The statistical pipeline comprised the following procedures:

1. Internal Consistency: Reliability of the Luria Raw Engine was calculated using Cronbach’s *alpha*, incorporating Bessel’s correction (*n* - 1) for unbiased population variance estimation.
2. Network Intercorrelations: Inter-domain relationships among the 10 cognitive indices and preset pathological network hubs (*preset_vci_svd, preset_msa, preset_ccas, preset_thalam, preset_retic, preset_striar*) were evaluated using Pearson correlation coefficients (Pearson *r*).
3. Discriminant Validity: Between-group separation across clinical mental status tiers was evaluated using non-parametric Kruskal-Wallis *H*-tests via SciPy, incorporating ties corrections for rank averaging across discrete score ranges.
4. Latent Structure Mapping: Latent dimensions of cognitive impairment were extracted via principal component analysis (PCA). The primary component (General Deficit Factor) was isolated using power iteration applied to the covariance matrix, yielding PC1 component loadings across all 10 domains.

### Statements and Declarations

#### 1. Ethics and Institutional Compliance Statement

This study conformed to the World Medical Association Declaration of Helsinki and was conducted as a clinical practice audit and internal quality improvement initiative approved by the Institutional Review Board of the participating clinical center. Primary bedside assessments and syndromic profiling were conducted by hospital staff during acute and subacute stroke phases alongside routine standard of care. Subsequent statistical analysis of the de-identified registry evaluated algorithmic precision without modifying clinical management, pharmacotherapy, or surgical interventions.

#### 2. Data Availability Statement

The primary de-identified numerical dataset, comprising the 10 Luria Raw domains, angiographic modifiers, and network hub presets, was completely de-identified in accordance with the HIPAA Safe Harbor method. De-identified data are available for academic research upon reasonable request to the corresponding author at Cognicore Neurosystems. Public repository deposition is restricted to safeguard proprietary algorithmic intellectual property and maintain institutional hospital database confidentiality.

## RESULTS

### Demographic and Variance Parameters of the Hospital Registry

The final statistical analysis included a cleaned cohort of *n* = 169 verified cases (*n* = 84 men, *n* = 85 women). The mean patient age was 66.87 years (*SD* = 13.9, range: 18.0–97.0 years). Variance parameters and cumulative cognitive deficit severity across four stratified age cohorts are summarized in Table 1. Mean cumulative deficit scores increased with age (from M = 5.08 in the young group to M = 9.24 in the advanced-age group). Marked sample size disparities across subgroups (*n* = 13 vs. *n* = 86) and significant between-group heteroscedasticity (variance shift *s*^2^from 8.41 to 45.57) rendered classical parametric analysis of variance (ANOVA) methodologically unstable. To maintain mathematical rigor, between-group differences were evaluated exclusively using non-parametric rank tests robust to unequal sample sizes and heterogeneity of within-group variances.

**Table 1.** Variance Parameters and Cognitive Deficit Across Age Cohorts.

| Age Cohort | n | Mean Total Deficit | 95% CI for Mean | Variance (s<sup>2</sup>) | SD |
| --- | --- | --- | --- | --- | --- |
| Young (<45) | 13 | 5.08 | 95% CI = [3.50; 6.65] | 8.41 | 2.90 |
| Middle (45-60) | 25 | 6.16 | 95% CI = [4.29; 8.03] | 22.72 | 4.77 |
| Elderly (61-75) | 86 | 7.43 | 95% CI = [6.00; 8.86] | 45.57 | 6.75 |
| Advanced (>75) | 45 | 9.24 | 95% CI = [7.55; 10.94] | 33.73 | 5.81 |

Sex-stratified analysis showed a mean total deficit of 6.91 points in men (*n* = 84) compared to 7.96 points in women (*n* = 85).

### Correlation Matrix: Expert Network Presets vs. 10 Luria Raw Domains

Intercorrelation analysis (Pearson r) evaluating associations between expert neural hub degradation presets and fractional scores across the 10 diagnostic domains is presented in Table 2.

**Table 2.**
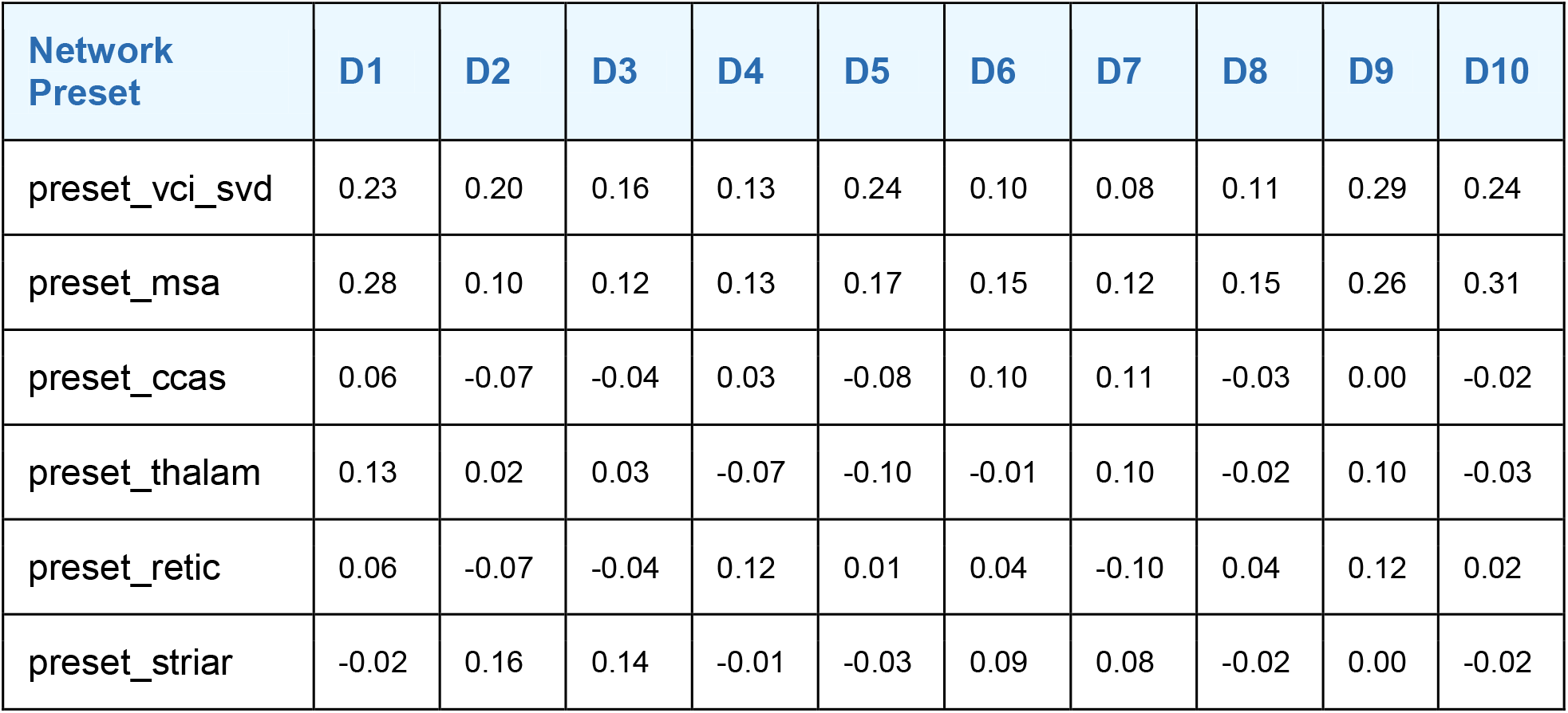
Pearson Linear Correlation Matrix (Network Presets vs. Domains)

Exploratory analysis of the cognitive-affective overlay demonstrated a linear Pearson correlation of r = -0.20 between cumulative cognitive deficit severity and depressive spectrum preset loading.

### Domain Discriminant Validation (Kruskal-Wallis Test)

Discriminant validity for each domain across multilevel mental status tiers (mental_status_code, k = 8 subgroups, total sample N = 169) was evaluated using the non-parametric Kruskal-Wallis H-test. In accordance with SAMPL guidelines, degrees of freedom were fixed at df = 7 (k - 1). Computations executed in SciPy incorporated tied rank averaging (ties correction) to account for discrete bounded score distributions. Statistical validation results are detailed in Table 3 (subgroup sample distributions across all eight categories appear in Table 4).

**Table 3.**
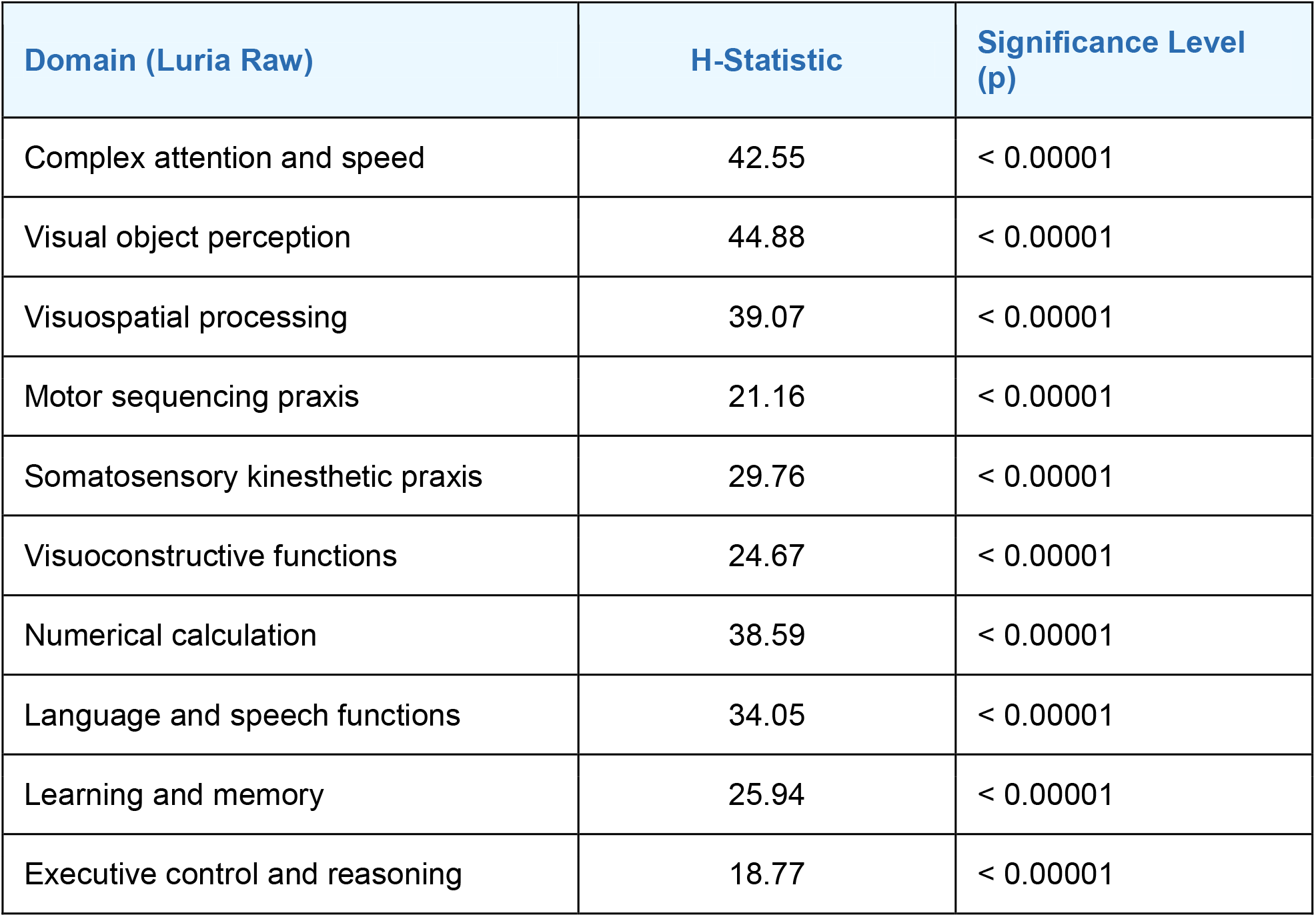
Discriminant Sensitivity of Individual Luria Raw Domains (SciPy Output)

| Domain (Luria Raw) | H-Statistic | Significance Level (p) |
| --- | --- | --- |
| Complex attention and speed | 42.55 | < 0.00001 |
| Visual object perception | 44.88 | < 0.00001 |
| Visuospatial processing | 39.07 | < 0.00001 |
| Motor sequencing praxis | 21.16 | < 0.00001 |
| Somatosensory kinesthetic praxis | 29.76 | < 0.00001 |
| Visuoconstructive functions | 24.67 | < 0.00001 |
| Numerical calculation | 38.59 | < 0.00001 |
| Language and speech functions | 34.05 | < 0.00001 |
| Learning and memory | 25.94 | < 0.00001 |
| Executive control and reasoning | 18.77 | < 0.00001 |

**Table 4.** Association Between Mental Status Codes and Total Deficit.

| Mental Status Code | Mean Total Deficit (M) | n | 95% CI for Mean |
| --- | --- | --- | --- |
| Code 7 | 1.01 | 6 | [-0.02; 2.04] |
| Code 0 | 5.56 | 102 | [4.72; 6.93] |
| Code 1 | 8.79 | 26 | [7.53; 10.06] |
| Code 0sleep | 9.00 | 10 | [6.79; 11.21] |
| Code 4 | 9.75 | 2 | [8.28; 11.21] |
| Code 2 | 13.98 | 16 | [10.49; 16.73] |
| Code 3 | 17.00 | 6 | [14.14; 19.86] |
| Code 5 | 20.00 | 1 | [0.00; 0.00] |

### Latent Structure and Empirical Validation of Mental Status Codes

Latent structure mapping via power iteration extraction of the covariance matrix’s leading eigenvector (*PC1*) established a deterministic hierarchy of post-stroke multidomain impairment. Dimensionality reduction to a single general deficit factor met Kaiser’s criterion: only the first principal component yielded an eigenvalue greater than unity (*λ*_*1*_= 4.1264), accounting for 41.26% of total feature variance. All subsequent components yielded eigenvalues *λ* < 1.0, reflecting statistical noise from comorbid and individual variance. Factor loadings across the 10 domains emerged in the following sequence:

- Visuoconstructive functions (D6): 0.46
- Language and speech functions (D8): 0.36
- Complex attention and speed (D1): 0.34
- Learning and memory (D9): 0.34
- Somatosensory kinesthetic praxis (D5): 0.32
- Motor sequencing praxis (D4): 0.28
- Executive control and reasoning (D10): 0.28
- Visuospatial processing (D3): 0.27
- Visual object perception (D2): 0.24
- Numerical calculation (D7): 0.21

Concordance between verified mental status codes and empirical mean cognitive deficit indices is detailed in Table 4. The Luria Raw Total Index (M) is calculated strictly as the additive sum of absolute penalty points across all 10 cognitive domains (theoretical range: 0 [normal baseline] to 50 [total functional collapse]), rather than through arithmetic or within-domain averaging, ensuring linear scalability of the composite index.

## DISCUSSION

### Interpretation of Latent Structure (PC1) and the Visuoconstructive Core Phenomenon

Principal component analysis (PCA) of the latent deficit structure reveals a fundamental neurobiological hierarchy underlying post-stroke multidomain impairment. Extracting the general deficit factor (*PC1*, eigenvalue *λ*_*1*_= 4.13, accounting for 41.26% of total feature variance) demonstrated that the visuoconstructive functions domain (D6) yielded the highest factor loading (*PC1* component loading = 0.46). This mathematical finding aligns with the classical theory of systemic dynamic localization of higher cortical functions. Within the framework of Luria’s syndromic analysis (Luria & Tsvetkova, 1966; Glozman, 1999), constructive praxis relies on the temporo-parieto-occipital (TPO) junction—a multimodal cortical hub that synthesizes spatial representations (Unit II) with action programming and executive sequencing (Unit III).

Following acute ischemic disruption, particularly cascading middle cerebral artery (MCA) syndromes, this metabolically demanding network node fails early, serving as the most sensitive indicator of generalized cerebral damage. In sharp contrast, numerical calculation (D7) exhibited distinct mathematical autonomy, contributing the lowest diagnostic weight to the general factor (PC1 Weight = 0.21). Basic arithmetic operations constitute overlearned, automated skills acquired early in ontogeny, rendering them resilient to generalized cognitive decline. Consequently, calculation breaks down primarily in circumscribed, focal lesions of the dominant parietal cortex (acalculia). The low factor loading of D7 alongside the elevated weight of D6 confirms the discriminant validity of the deterministic core engine: rather than redundantly sampling shared variance, the platform’s domains demonstrate strict orthogonality, measuring clinically and anatomically distinct neural networks.

### Neural Network Decoding of Expert Presets

Intercorrelation analysis of the network hub matrix translates classic clinical syndromes into quantitative parameters. The cerebral microangiopathy profile (preset_vci_svd) correlated strongest with memory (D9 = 0.29), kinesthetic praxis (D5 = 0.24), and complex attention (D1 = 0.23). This constellation reflects the classical disconnection syndrome in network neuroscience (Mesulam, 1990; Seeley et al., 2007). Deep white matter pathology in small vessel disease (SVD) disrupts corticosubcortical loops, primarily compromising processing speed and mnestic retrieval dynamics while sparing core cortical nodes (Peña-Casanova et al., 2006). This topographical dissociation is evident in Figure 1, where anterior circulation infarctions (carotid territory, pool = 1) produce peak deficits in kinesthetic praxis and speech/language domains. The cerebellar preset (preset_ccas) showed selective orthogonality. The deterministic engine captured near-zero or negative correlations with canonical cortical domains: speech/language (D8 = -0.03), object gnosis (D2 = -0.07), and executive functions (D10 = -0.02). Conversely, positive associations emerged with numerical calculation (D7 = 0.11) and visuoconstructive praxis (D6 = 0.10). These data provide computational verification of cerebellar cognitive dysmetria (Schmahmann & Sherman, 1998). The selective involvement of cerebello-brainstem pathways alongside spared visual domain functioning is evident in Figure 1: the vertebrobasilar cohort (pool = 2) demonstrated preserved visual object perception (0.33) despite marked deficits in complex attention and constructive functions. The cerebellum does not generate lexical semantics or primary perceptual representations; rather, it calibrates the spatiotemporal coordination and metric precision of cognitive operations via reciprocal cerebello-cerebral loops. Subcortical analysis revealed distinct contributions from thalamic (preset_thalam) and reticular (preset_retic) presets, indexing dysfunction within Unit I (arousal and neurodynamics). Both presets primarily affected learning and memory dynamics (D9 = 0.10–0.12). However, motor sequencing exhibited divergent loadings: the reticular pattern impaired motor set-shifting fluency (D4 = 0.12), whereas the thalamic pattern showed a negative association (D4 = -0.07), reflecting the fluctuating, asthenic profile of subcortical deficits. Learning & Memory represented the point of absolute convergence across both vascular pools (Figure 1: 1.84 vs. 1.82), confirming that amnestic deficit is an obligatory feature of acute stroke regardless of primary lesion site.

**Figure 1.**
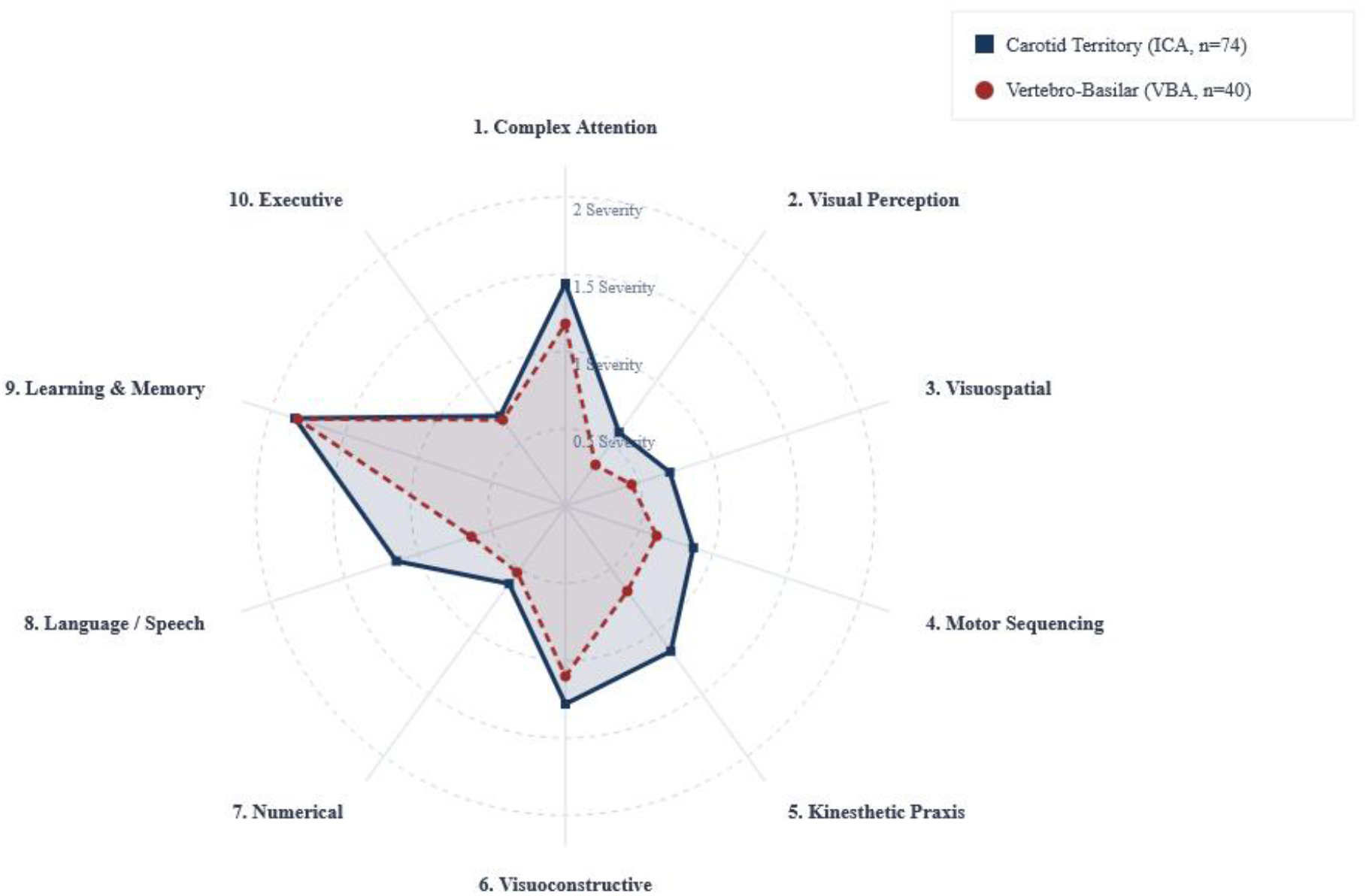
Topographical Neurocognitive Deficit Profiles Across Vascular Pools (n = 114 Processed Acute Stroke Records). Note: The topographical profile visualization (Figure 1) is derived from a targeted subanalysis of the vascular cohort (n = 114). The analysis excluded 55 records categorized in Table A as clinical_pool = 0 (n = 54 somatic controls lacking focal vascular lesions; n = 1 acute stroke case with incomplete initial vascular localization). The final profile directly compares patients with carotid circulation occlusions (n = 74, pool = 1) against vertebrobasilar circulation occlusions (n = 40, pool = 2). Higher values along the radial axes indicate greater deficit severity on the continuous Luria Raw metric (0 to 5).

### Dissociation of Pseudodementia and Validation of Mental Status Codes

The weak negative Pearson correlation between cumulative cognitive deficit severity and depressive preset loading (*r* = -0.20) rules out a clinically significant contribution of affective pseudodementia to the general deficit factor. This coefficient reflects a measurement ceiling rather than a direct psychopathological mechanism: severe multidomain collapse (high Luria Raw Total Index) introduces executive dysfunction and aphasic barriers that preclude valid self-report screening—a recognized validation failure of self-report scales in acute neurological cohorts. In the absence of severity-stratified affective screening, this correlation confirms that the cognitive measurement core operates independently of transient acute emotional fluctuations. The monotonic gradient observed across admission mental status tiers—with mean total deficit scores rising from *M* = 1.01 (Code 7) to *M* = 20.00 (Code 5)—confirms the clinical discriminant validity of this intake classification. Rather than imposing a rigid linear severity metric, these codes serve as deterministic operational presets documenting verbal rapport, cooperation, and affective tone at the time of bedside assessment. Clinicians assign these codes during initial intake: patients with intact rapport and euphoric or anosognosic tone (frequent in right-hemisphere stroke) receive baseline Code 0 (*M* = 5.56), whereas isolated emotional lability (Code 7, *M* = 1.01) and transient neurodynamic slowing secondary to sedation or sleep deprivation (Code 0sleep, *M* = 9.00) are isolated systematically. As behavioral disorganization worsens across codes, the cumulative quantified deficit scales accordingly. Crucially, the platform scores the 10 cognitive domains independently through objective performance on standardized test paradigms rather than behavioral observation. Decoupling behavioral rapport from cognitive testing prevents diagnostic artifacts, anchoring intake observations directly to objective neural network profiles.

### Study Limitations and Future Directions

Despite the granular clinical characterization of the sample, several limitations must be acknowledged. First, the investigation utilized a retrospective observational design based on registry data. Second, the 7-month recruitment window excluded rare vascular syndromes, notably pure callosal disconnection syndromes, due to the extremely low clinical incidence of isolated pericallosal artery occlusions in unselected emergency admissions.

Looking forward, this computational core enables prospective tracking of post-stroke cognitive recovery. The deterministic framework may be adapted for longitudinal monitoring in rehabilitation settings to quantify cognitive recovery trajectories objectively.

## CONCLUSION

This study validates a methodological framework for quantifying Luria’s qualitative syndromic analysis and the Boston Process Approach within acute clinical workflows. Converting semiotic indicators of higher cortical dysfunction into continuous fractional metrics (Luria Raw) formalizes neurocognitive deficit profiles, mitigating subjective examiner bias and scoring variability inherent to traditional manual recording. Extracting the first principal component as an integrated cognitive deficit index (PC1), combined with the verified statistical independence of individual cognitive domains, establishes a robust measurement foundation for objectively assessing the structure and evolution of post-stroke multidomain impairments during acute cerebral ischemia.

A second major contribution is the clinical validation of a two-tier data processing pipeline to ensure diagnostic reliability. Pinning primary semiotic markers from syndromic analysis within an isolated deterministic measurement core engine prior to text generation resolves a central barrier to clinical large language model (LLM) integration: algorithmic vulnerability to stochastic drift and stochastic errors. Restricting generative models strictly to last-mile linguistic synthesis within a bounded tabular context eliminates clinical fact distortion. This architecture guarantees report auditability and aligns with the architectural requirements of client-side HIPAA and GDPR data governance standards.

Clinical implementation of this paradigm standardizes diagnostic workflows across interdisciplinary medical settings. In multicenter clinical trials, automated high-resolution profiling eliminates inter-rater reliability bias, ensuring reproducible tracking of deterministic deficit trajectories. In academic medical centers, this standardization automates the export of structured datasets directly compatible with multivariate statistical pipelines in SciPy, R, or SPSS. Finally, automated mapping of deficits to ICF categories provides standardized functional metrics to assist rehabilitation planning in acute care units.

## Notes

### Competing Interest Statement

The corresponding author of this study is the founder and lead developer of Cognicore Neurosystems, the technology group developing the NeuroDraft software platform evaluated in this study. This commercial interest did not influence the objective statistical processing, data analysis, or reporting of the clinical registry results presented herein.

### Author Declarations

Ethics committee and Institutional Review Board of Botkin City Clinical Hospital gave ethical approval for this work.

